# A Discernable Increase in the Severe Acute Respiratory Syndrome Coronavirus 2 R.1 Lineage Carrying an E484K Spike Protein Mutation in Japan

**DOI:** 10.1101/2021.04.04.21254749

**Authors:** Tsuyoshi Sekizuka, Kentaro Itokawa, Masanori Hashino, Kazuhiro Okubo, Asami Ohnishi, Keiko Goto, Hiroyuki Tsukagoshi, Hayato Ehara, Ryohei Nomoto, Makoto Ohnishi, Makoto Kuroda, Virus Diagnosis Group (NIID Toyama) and COVID-19 Genomic Surveillance Network in Japan (COG-JP).

## Abstract

Three COVID-19 waves in Japan have been characterized by the presence of distinct PANGO lineages (B.1.1. 162, B.1.1.284, and B.1.1.214). Recently, in addition to the B.1.1.7 lineage, which shows 25% abundance, an R.1 lineage carrying the E484K mutation in the spike protein was found to show up to 40% predominance.

## Text

As of March 31, 2021, Japan reported 475,043 cases and 9,175 deaths due to coronavirus disease (COVID-19), some of which were caused by variants of concern (VOCs). We conducted genome surveillance for severe acute respiratory syndrome coronavirus 2 (SARS-CoV-2) with support from local public health centers or laboratory institutes (*1,2*) and five airport quarantine stations (Narita, Haneda, Nagoya, Kansai, and Fukuoka airports) (*3*). During the surveillance, we monitored several VOCs of the PANGO lineage, namely B.1.1.7 (501Y.V1), B.1.351 (501Y.V2), and P.1 (501Y.V3) (*4*), which have emerged in the past months in all countries. As of March 31, 2021, a notable increase in imported VOC cases has been identified at the airport (123 cases) and domestic (678 cases) quarantines. COG-JP has been monitoring the prevalence of PANGO lineages consistently, from the first COVID-19 case (January 15, 2021) up to recent cases (March 6, 2021) [≥29 kb genome in size; in total, 28,350 isolates have been deposited in the Global Initiative on Sharing All Influenza Data (GISAID) EpiCoV database (Appendix Table 1)]. The study protocol was approved by the National Institute of Infectious Diseases, Japan (approval no. 1091). The ethics committee waived the written consent requirement for research on the viral genome sequence.

We have found notable distribution of the PANGO lineage during each COVID-19 wave. Specifically, B.1.1.162, B.1.1.284, and B.1.1.214 were predominant during the first, second, and third waves, respectively (Figure 1). After the first wave (March to April 2020), an effort to monitor airport quarantine was initiated; this measure was successful in reducing the numbers of imported cases (3). Domestic lineages (B.1.1.284 and B.1.1.214), which may have been derived phylogenetically from B.1.1.162 during the first wave, were predominant during the second wave (July to September 2020). However, this was not the case for the other imported lineages. Moreover, B.1.1.214 was the predominant lineage during the peak of the third wave (October 2020 to January 2021) (Figure 1).

**Figure 1.**
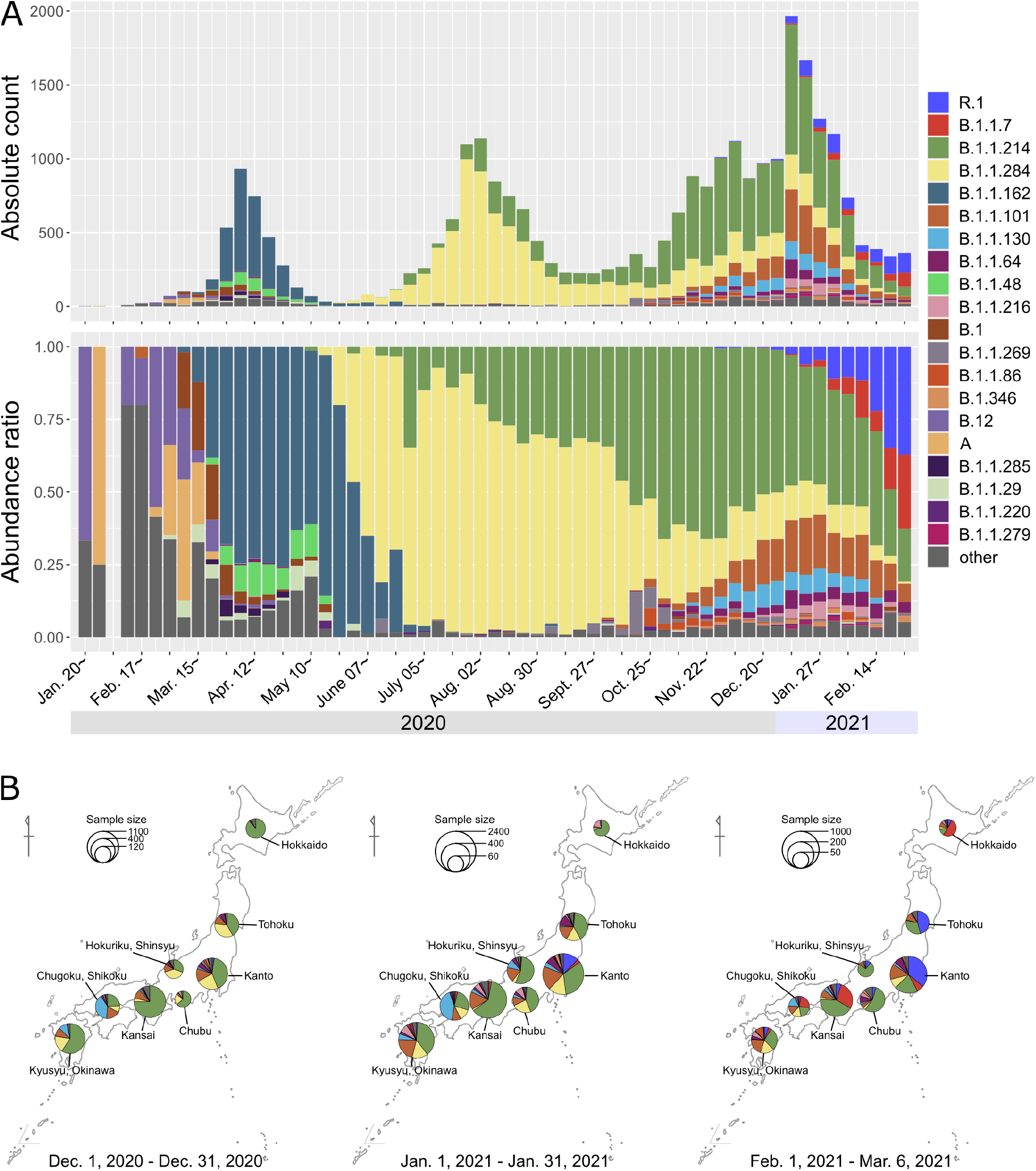
A spatiotemporal analysis of the SARS-CoV-2 PANGO lineage in Japan. A) Numbers and B) abundance ratios of whole-genome sequences of SARS-CoV-2 isolates from domestic COVID-19-positive patients in Japan (n = 28,965) were assigned as per the PANGO lineage definition (2021/02/21 version). Top 20 PANGO lineages have been highlighted as per the colors on the right. C) Area-specific lineage percentage is displayed using colored pie charts (shown in panel A and B) from December 2020 to February 2021.

Recently, a new R.1 lineage carrying the spike protein mutation E484K has been detected in < 5% to > 37.6% of all COVID-19-positive cases; within a span of six weeks (mid-January to early March 2021), there has been a sharp increase in the cases (Figure 1). The spatiotemporal distribution of the PANGO lineage showed that the R.1 cases appear to be predominant in the Kanto and the northern Tohoku area. Our study, therefore, highlights the region-specific transmission of COVID-19. Indeed, B.1.1.7 cases have been especially predominant in the Kansai area; however, data from the ongoing surveillance of other VOCs will be required to confirm the extensive transmission of B.1.1.7 across Japan.

As a notable feature of VOCs, the B.1.1.7 variant is susceptible to neutralizing antibodies elicited by vaccines using the ancestral spike protein (5). However, there is a greater concern about the other immune evasion mutations, such as the E484K (Glu484→Lys) mutation in the spike (S) protein that is found in the B.1.351 (501Y.V2) variant that emerged in South Africa and the P.1 (501Y.V3) variant in Brazil (6). Indeed, E484K could be a pivotal amino acid substitution with potential for mediating immune escape. Thus far, B.1.1.318, B.1.525, R.1, R.2, and P.2 have been reported as variants carrying the E484K single mutation on the receptor-binding domain. R.1 has been mostly identified in USA (first found on October 24, 2020) and Japan (first found on November 30, 2020). In fact, phylogenetic analysis indicated that both R.1 isolates were highly similar. Thus, the Japanese R.1 isolate might have originated from the R.1 isolate from USA. Conversely, it is possible that the isolates in both countries have been imported from an uncharacterized source in another country, where a potential common ancestor (B.1.1.316) was circulating (Appendix Figure 1).

In conclusion, although VOCs have a marked impact on the number of cases and the severity of COVID-19 across many countries, mutants of concern carrying pivotal mutations should not be disregarded if the latent distribution has been found across the community. We should pay more attention to such potential VOCs to avoid the emergence of mutants of concern. Based on the current epidemiological situation in Japan, with an increase in the circulation of more transmissible lineages, immediate, strong, and decisive public health interventions such as contact-tracing, strict quarantine monitoring, and comprehensive real-time genome surveillance have become essential for containment of COVID-19 clusters.

## Supporting information

Appendix Table 1_GISAID

Appendix Table 2_Collaboration-COG-JP

## Data Availability

COG-JP has been monitoring the prevalence of PANGO lineages consistently, from the first COVID-19 case (January 15, 2021) up to recent cases (March 6, 2021) [≥29 kb genome in size; in total, 28,350 isolates have been deposited in the Global Initiative on Sharing All Influenza Data (GISAID) EpiCoV database (Appendix Table 1)]. We would like to thank all researchers who deposited and shared genomic data on GISAID (Column H-J in Appendix Table 1).

## Acknowledgements

This work was supported by a Grant-in-Aid from the Japan Agency for Medical Research, and the Development Research Program on Emerging and Re-emerging Infectious Diseases [JP20fk0108103 and JP19fk0108104, respectively]. We would like to thank all researchers who deposited and shared genomic data on GISAID (Column H-J in Appendix Table 1). The collaborators are listed in Appendix Table 2.

## Author Bio

Dr. Sekizuka is the chief of the Pathogen Genomics Center, National Institute of Infectious Diseases, Japan. He developed bioinformatics tools to elucidate the transmission dynamics, surveillance, and control of infectious diseases based on pathogen genomics.

---

**Appendix Table 1**.

Collaboration with local public health institutes and airport quarantines under the COG-JP have been indicated. All SARS-CoV-2 complete genome sequences that we determined and other deposited sequences identified in Japan were retrieved from the GISAID (n = 28,965). COG-JP, COVID-19 genomic surveillance network in Japan; GISAID, Global Initiative on Sharing All Influenza Data; SARS-CoV-2, severe acute respiratory syndrome coronavirus 2

**Appendix Table 2**.

Collaboration between local public health institutes and the COVID-19 genomic surveillance network in Japan (COG-JP) has been shown.

**Appendix Figure 1.**
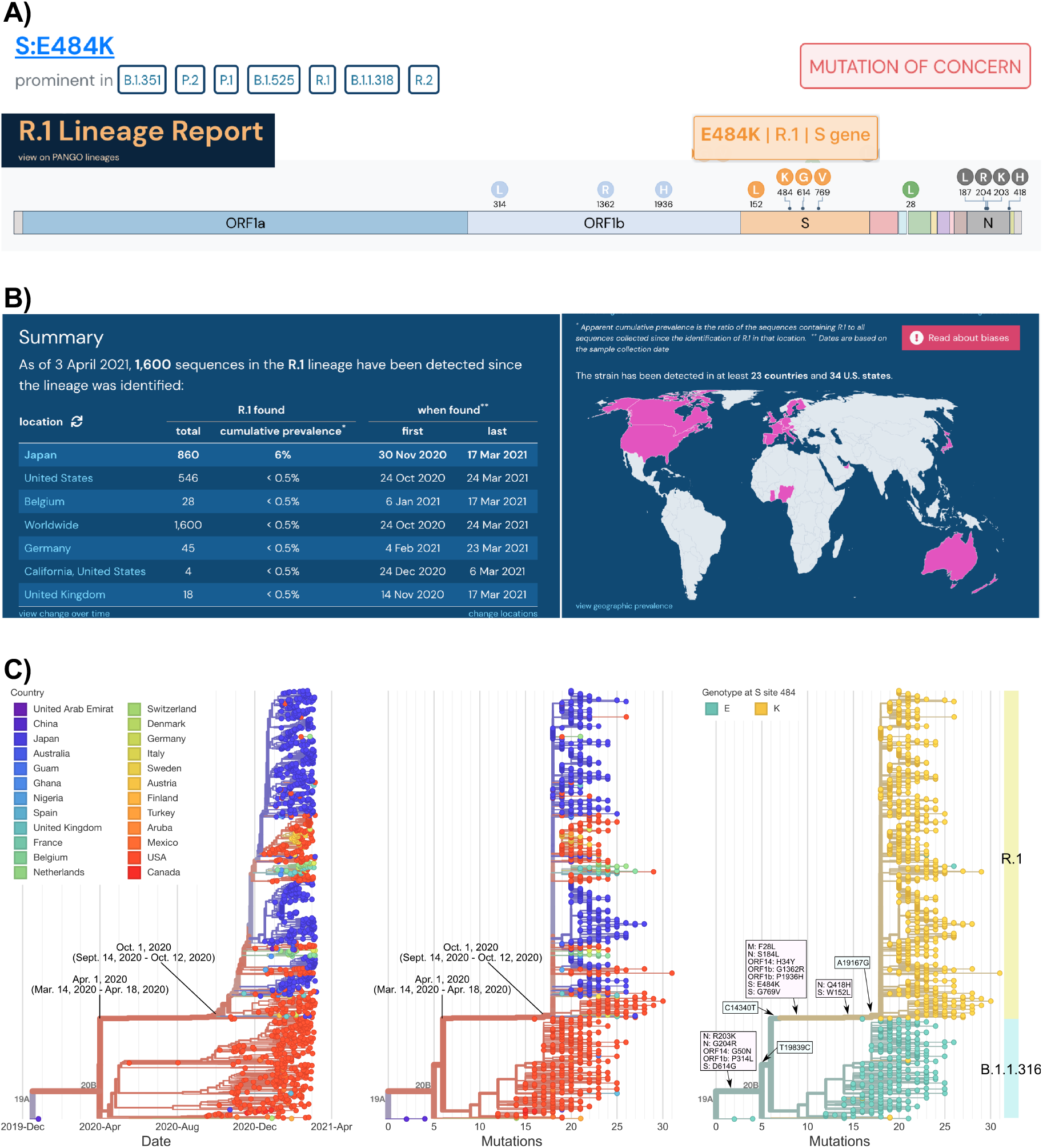
Summary of SARS-CoV-2 R.1 lineage (April 3^rd^, 2021) A) The R.1 lineage has a mutation of concern. The R.1 lineage carries the E484K mutation in the S protein; common genetic features are highlighted above the schematic representation of the SARS-CoV-2 genome structure. B) Current information on the number of cases, country, and first detection date for the R.1 lineage (7). C) Phylogenetic analysis of the R.1 lineage by Nextstrain analysis, along with the B.1.1.316 lineage which is the most recent common ancestor (8).

